# Knowledge mobilization activities to support decision-making by youth, parents, and adults using a systematic and living map of evidence and recommendations on COVID-19: protocol for three randomized controlled trials and qualitative user-experience studies

**DOI:** 10.1101/2022.05.09.22274842

**Authors:** Rana Charide, Lisa Stallwood, Matthew Munan, Shahab Sayfi, Lisa Hartling, Nancy J. Butcher, Martin Offringa, Sarah Elliott, Dawn P. Richards, Joseph L. Mathew, Elie A. Akl, Tamara Kredo, Lawrence Mbuagbaw, Ashley Motillal, Ami Baba, Matthew Prebeg, Jacqueline Relihan, Shannon D. Scott, Jozef Suvada, Maicon Falavigna, Miloslav Klugar, Tamara Lotfi, Adrienne Stevens, Kevin Pottie, Holger J. Schünemann

## Abstract

**Introduction:** The COVID-19 pandemic underlined that guidelines and recommendations must be made more accessible and more understandable to the general public, including adults, parents, and youth, to improve health outcomes. The objective of this study is to evaluate, quantify, and compare the public’s (youth, parents, and adult populations) understanding, usability, satisfaction, intention to implement, and preference for different ways of presenting COVID-19 health recommendations derived from the COVID-19 Living Map of Recommendations and Gateway to Contextualization (herein referred to as the RecMap).

**Methods and Analysis:** This is a protocol for a multi-method study. We will conduct pragmatic allocation-concealed, blinded superiority randomized controlled trials (RCT) in three populations to test alternative formats of presenting health recommendations: adults (21 years of age or older), parents (18 years or above and are a parent or legal guardian of a child under 18 years old), and youth (15 to 24 years old), with at least 240 participants in each population. The research will consist of a randomized online survey and an optional one-on-one interview. Prior to initiating the RCT, our interventions will have been refined with relevant stakeholder input. In each population group, the intervention arm will receive a plain language recommendation (PLR) format while the control arm will receive the corresponding original recommendation format as originally published by the guideline organizations (herein referred to as Standard Language Version). Our primary outcome is understanding, and our secondary outcomes are accessibility and usability, satisfaction, intended behavior, and preference for the two recommendation formats. Each population’s results will be analyzed separately. However, we are planning a meta-analysis of the results across populations, and will also explore potential interaction and subgroup effects within each population. At the end of each survey, participants will be invited to participate in a one-on-one, virtual semi-structured interview to explore their user experience and their learning preferences and future research. All interviews will be transcribed and analyzed using the principles of thematic analysis and a hybrid inductive and deductive approach. Iterative member checking, triangulation, interpretation, and saturation of themes will be sought to enhance reliability.

**Ethics and Dissemination:** Through Clinical Trials Ontario (CTO), the Hamilton Integrated Research Ethics Board has reviewed and approved this protocol (Project ID: 3856). The University of Alberta has approved the parent portion of the trial (Project ID:00114894). All potential participants will be required to provide informed consent. The findings from this study will be disseminated through open-access publications in peer-reviewed journals and using social media.

**Strengths and limitations of this study:**

- We are following a multi-method approach: randomized controlled trials and qualitative interviews. The qualitative results will supplement and help explain our quantitative findings.
- This protocol is reported in accordance with the Standard Protocol Items: Recommendations for Interventional Trials (SPIRIT), which enhances transparency and completeness. The trials use previously validated outcomes from similar trials. This will strengthen the credibility of our results.
- Our study is testing an optimized plain language recommendation format, which makes our intervention relevant to our stakeholder groups, and is recruiting internationally, which ensures the inclusion of a diverse population. Recruitment will take place online using social media, and data will be collected using an online survey. This allows for self-selection and limits accessibility to those who have no or limited digital access, which in turn limits generalizability.
- While the recommendations are offered in multiple languages through the RecMap, the study is only testing English plain language recommendation summaries.

## Introduction

As health practitioners, policymakers, and the public are inundated with information, misinformation, and health recommendations about COVID-19, we have built a unique repository of trustworthy COVID-19 recommendations with funding from the Canadian Institutes of Health Research (CIHR) (covid19.recmap.org). The overarching goal of this COVID-19 RecMap effort is to identify all COVID-19 guidelines, assess their credibility and trustworthiness, and make their recommendations available and understandable to various stakeholder groups [1].

To enhance understanding of health recommendations by the public, we developed a multi-stakeholder process to draft, edit, and publish plain language recommendations (PLRs) [2]. Until now, plain language versions of recommendations have been poorly explored, and our work on the RecMap indicates an absence of this critical knowledge mobilization tool for the general public, including youth, adults, and parents [3]. Existing trials on tailored recommendations presentations have been small and predated recent guidance on how to present guideline recommendations, targeted health professionals, or did not target the general public [4-7]. For example, a small trial with 84 mental health service users suggested improved intention to follow recommendations when written in plain language [6]. Trial data also exists on specific aspects related to health information, like comparing alternative ways of presenting numerical or statistical information and formats of information sharing [4, 8-10]. However, trustworthy and comprehensive PLRs may need to include the clinical or public health background related to the topic, conflicts of interest, available research evidence, judgments that are made, the rationale for a recommendation, the actual recommendation, and implementation considerations. As addressed in Evidence to Decision (EtD) frameworks, these facets are deemed essential to inform decision-making and are used widely by various organizations [11-14].

A trial is necessary to empirically show that PLRs convey the intended messages from guidelines to broader populations and not only to the selected user groups that usually participate in ours and others’ research. Understanding and interpreting recommendations correctly are the essential prerequisites for the general public to become effective self-managers of their health and to ultimately optimize behaviors and related health outcomes [15]. Creating health information for the public that is accessible, reliable, and understandable is critical to scale science and evidence-based guidelines, which is especially important during the COVID-19 pandemic. We plan to undertake this trial, given the paucity of empirical evidence to guide our team and guideline developers as to how to best present PLRs and EtD facets to the public, which are critical to support their informed decision-making. This protocol describes the trials and details of the three leading trial sites.

## Objectives

The objective of this study is to compare end-users’ (youth, parents, and adult populations) understanding, accessibility and usability, satisfaction, intention to implement, and preference for COVID-19 recommendations when presented as plain language recommendation summaries (PLRs) versus standard language versions (SLVs). We hypothesize that there will be a difference in understanding of information between the two formats. We also aim to understand the reasons for their choices through our linked qualitative research.

## Methods

We will follow a multi-methods approach: randomized controlled trials (RCT) with qualitative interviews among a subset of participants. The trials are pragmatic allocation-concealed, blinded superiority RCTs with at least 240 participants in each population (parents, adult, and youth). The trials were preceded by preparatory engagement work with Canadian youth advisors, a Canadian parent advisory group, and an international adult Cochrane Consumer group, herein referred to as stakeholder groups. The youth site’s preparatory engagement work was co-developed and implemented by two youth partners (MP and JR) in collaboration with LS, to ensure meaningful engagement efforts were embedded into the research process. We gathered input from stakeholders on COVID-19 priority topics and PLR content and format. Input from stakeholders helped us refine the PLR format to be evaluated with each population in the trial.

We will conduct qualitative interviews with a subset of the participants who completed the trials to help contextualize the digital user experience and understand the results of the trial.

Any amendments to this protocol will be made available via medRxiv.

### Randomized Controlled Trials

This protocol has been prepared in accordance with the Standard Protocol Items: Recommendations for Interventional Trials (SPIRIT) reporting guideline (Appendix 1) [16].

### Design and Setting

There are three online survey links, one for each respective population. Each link contains eligibility questions, the consent form, and study information for youth, parents, or adults. Participants will access the online survey corresponding to their respective population to enter the trials and provide implied consent by participating in the survey. Using the concealed allocation code of the intervention platform, SurveyMonkey (surveymonkey.com), participants will select their geographic region. Participants from each geographic region will be randomized to one of two recommendation topics, and then they will be randomized to a PLR format (intervention) or a SLV format (control) of the recommendation (Figure 1). The SLV is the original PDF guideline developed and published by an organization, such as the World Health Organization, from where the PLR information was extracted. Participants will complete an online survey consisting of Likert-scale questions, multiple-choice questions, open-ended questions, and demographic information. The survey is anonymous.

**Figure 1.**
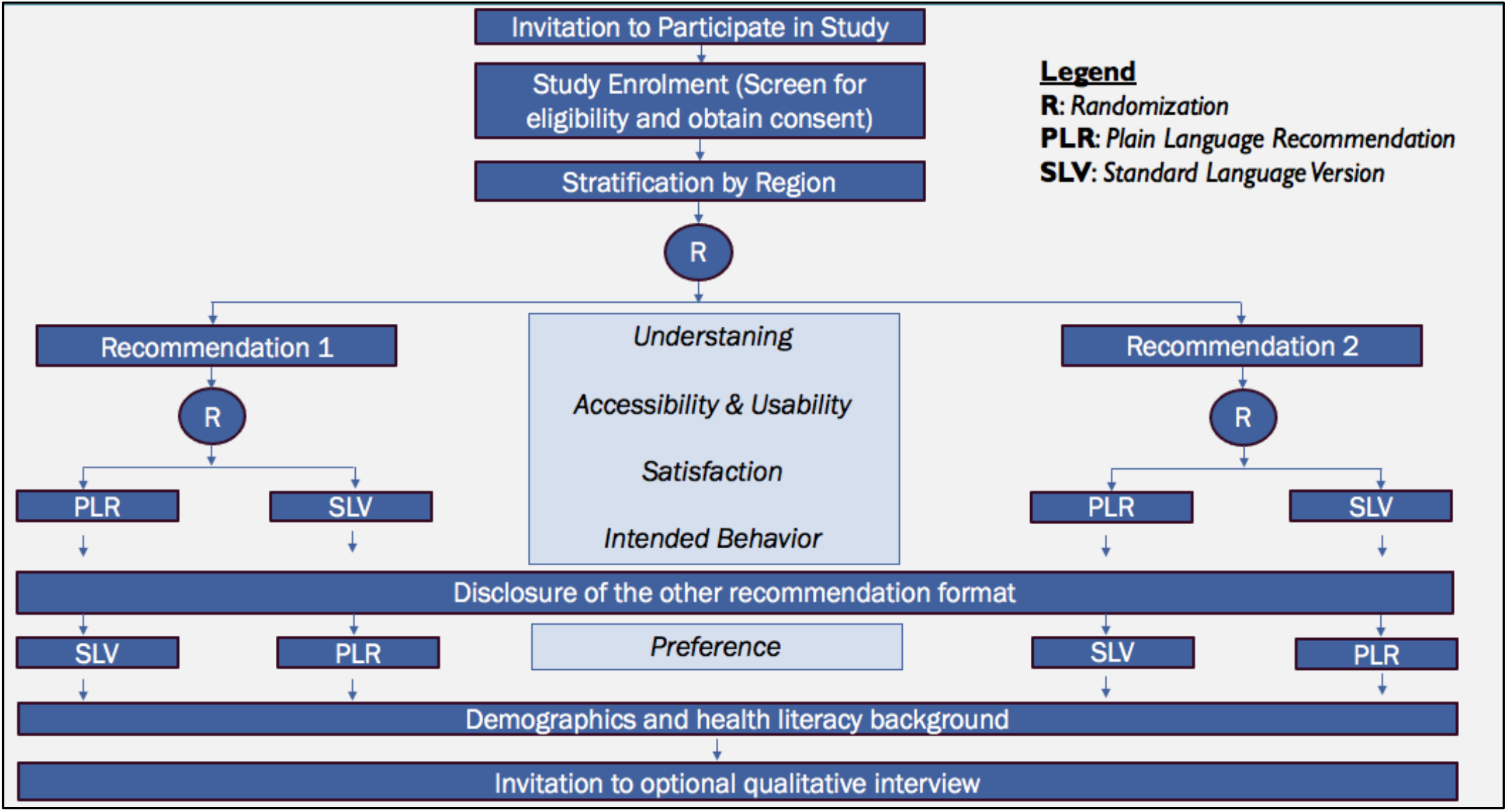
RCT Flow Diagram

Following completion of the online survey, we will invite participants to take part in an optional qualitative interview that will allow them to reflect on their preference for the PLR or SLV format and help us understand the results of the quantitative survey. Confidentiality will be maintained by storing names and emails on a password protected device and secure network to which only authorized study personnel have access to. If participants decide to provide their name and email address to participate in the interview, receive the study results, or enter the draw (for a chance to win one out of five $50 CAD gift cards for the adult and parents’ sites, one out of ten $25 CAD gift cards for the youth site) this personal information will not be linked to their survey answers.

This is not a clinical trial, thus there are no foreseeable harms or health risks from participating. However, there is a possibility that participants may experience distress once they better understand the uncertainty in the underlying evidence, even if the recommendation itself is judged trustworthy. We provide the email address and contact information of the principal investigator and research coordinator. Participants can contact us to report any discomfort or seek further information.

### Participants

Each of the three leading sites will recruit participants from one of the three population groups: adults, parents or youth. SickKids Research Institute will lead the recruitment of youth, Western University will lead the recruitment of adults, and The University of Alberta will lead the recruitment of parents. Global recruitment efforts will take place via social media. Coordination and oversight of the trials rests with the research team at McMaster University (RC and HJS). Our study is also open for other sites who are interested in participating as active recruitment sites for one or more populations. For example, St. Elizabeth University in Slovakia is joining as an active recruitment site for all three populations, with a commitment to recruit up to 60 participants.

### Selection Criteria

In this study, adults are defined as individuals self-reported to be 21 years of age or older, youth are defined as individuals self-reported to be between the ages of 15 to 24 years [17], and parents are defined as individuals self-reported to be 18 years or above and are a parent or legal guardian of a child under 18 years old. Participants can be from any country. For all three populations, participants will need to be able to read and understand English. We expect variation in the English language proficiency and health literacy level among participants. Therefore, when collecting demographic information, questions related to language proficiency and familiarity with COVID-19 health information will be included to understand the participant’s level of health literacy. We may adjust for these variables in our analysis if needed, although we expect them to be reasonably balanced between the SLV and PLR groups due to randomization.

### Recruitment

Each leading site will run recruitment for the different population groups separately yet with similar and overlapping strategies. We will recruit English-speaking (mother tongue or fluent) participants globally through our experienced RecMap investigator team (https://covid19.recmap.org/about), the Cochrane Consumer Network and various Cochrane networks, guideline co-authors who interact with youth (including the international Young Persons’ Advisory Groups), parents, and adult citizens. We plan to run public recruitment campaigns through identified youth, adult, and parent organizations’ social media platforms (e.g., organizations’ accounts on Twitter, Instagram, and Facebook) and newsletters. Each leading site will be responsible for monitoring their own recruitment, though all investigators will support overall recruitment for the trials (e.g., if recruitment falls behind for one of the populations, other sites will help support recruitment for that group). Surveys will be administered through SurveyMonkey, where participants are screened for eligibility prior to allocation. Previous studies suggest that the target recruitment rates for such trials are achievable in less than three months, and we will recruit at least 240 participants per group [18-20].

The average survey completion time is estimated to be 15-25 minutes, based on stakeholder feedback and pilot testing. As suggested by our stakeholders in the preparation of this trial, we will be inviting participants who complete the survey to enter a draw for a chance to win one of five $50 CAD gift cards for the adult and parents’ populations. The youth group will invite youth participants to enter a draw for one of ten gift cards valued at $25 CAD. This is offered as a gesture and appreciation for completing the survey.

### Intervention and Comparison

In these trials, the intervention is the PLR, and the comparison is the SLV. Figure 2 below shows an example of the SLV format of a recommendation that will be used with the adult and youth participants and Figure 3 shows an example of a PLR format that will be presented to adult participants.

**Figure 2.**
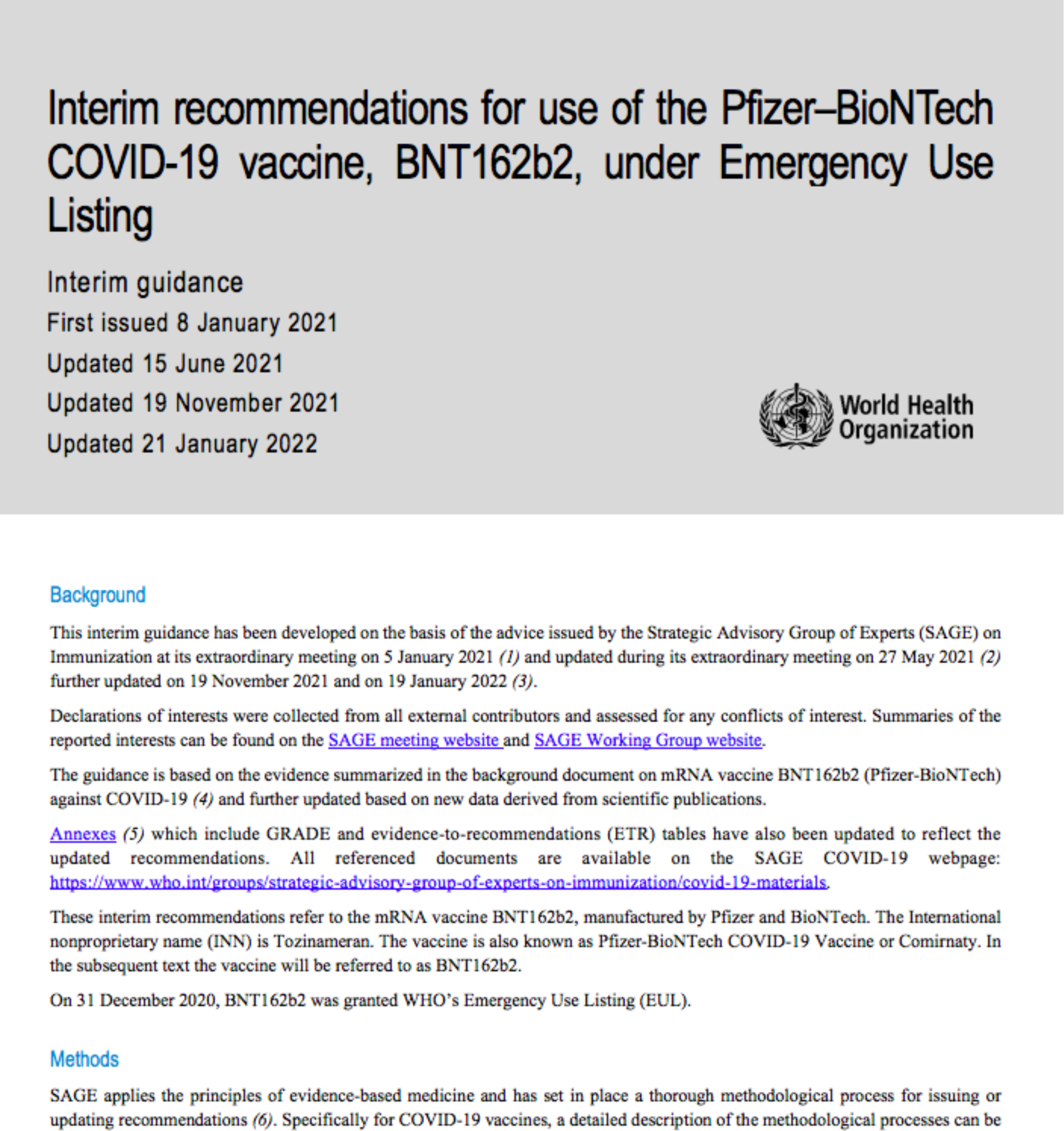
SLV Format for Youth and Adult Participants [21]

**Figure 3.**
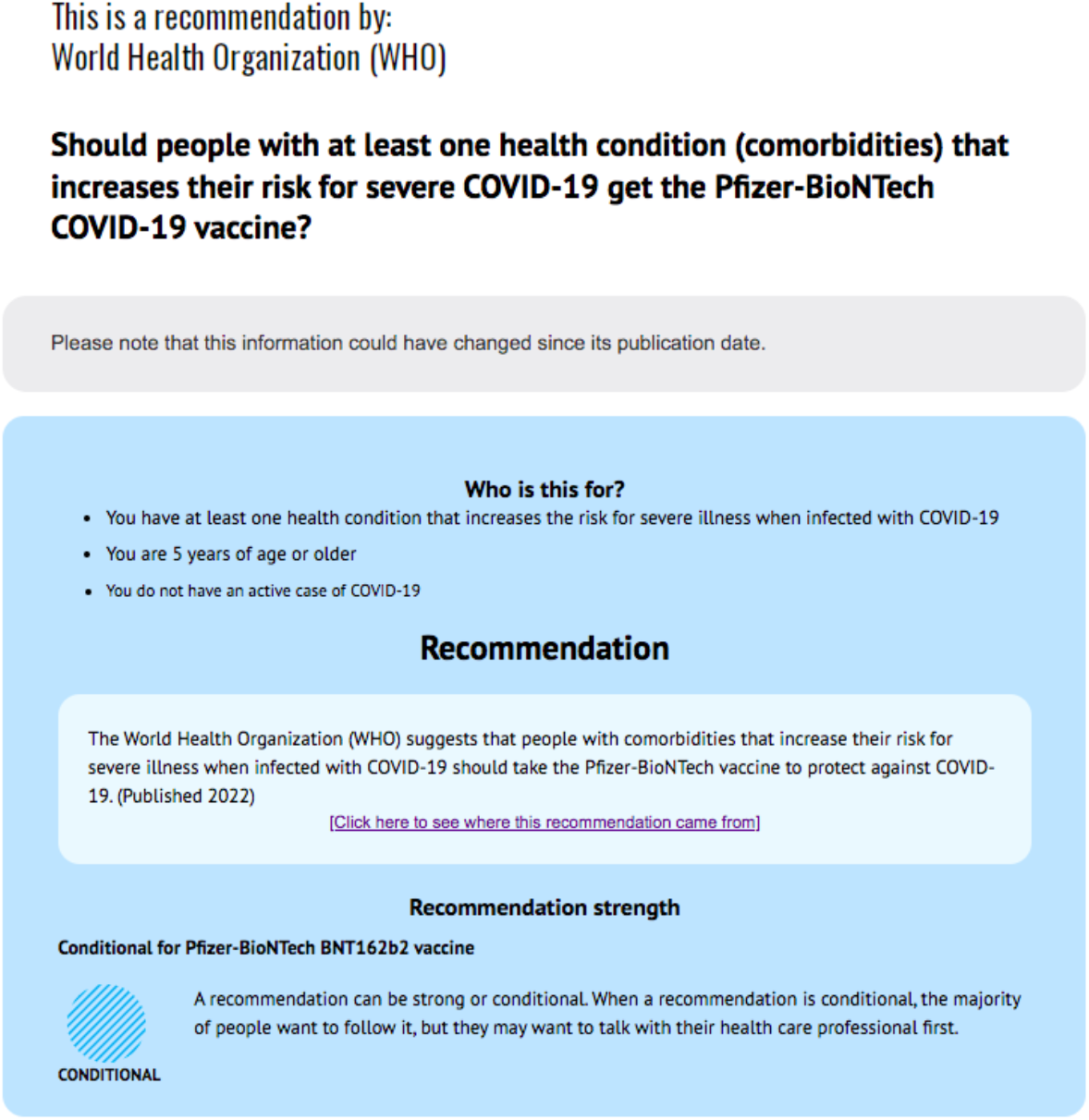
Example of a Plain Language Recommendation Summary Format for Adult Participants

Through our pre-trial engagement work, we gathered input from youth (via online workshops) and from adult stakeholders (via online meetings) on high priority COVID-19 topics and on PLR content and format. Input from youth and adult stakeholders allowed us to choose two recommendations for the trials and refine the PLR format. For the parents’ population, the PLRs were refined from collected feedback during online meetings with parent groups.

All PLRs for the trials were developed from the same template and refined using stakeholder feedback. The youth and parent PLRs’ design and formatting are the same, as these groups gave similar feedback, while the adult PLRs are slightly different.

In the trials, we will ask participants to read one format of a recommendation (PLR or SLV) to which they have been randomly allocated and then answer the online survey that takes an average of 15-25 minutes to complete.

### Outcomes

#### Primary Outcome

##### Understanding

We define understanding as the comprehension of key guideline content (e.g., year of publication, intent of recommendation, direction of recommendation, etc.). This outcome will be measured using seven multiple-choice questions about key concepts in the recommendation with four to six response options for each question and only one correct answer (total minimum score of 0, maximum score of 7). Based on previous work, we aim to detect a difference of 10% in understanding between groups which we consider an important difference (an average difference of 1 correct answer) [18, 19, 22-24].

#### Secondary Outcomes

##### Accessibility and Usability

We define accessibility and usability as the ability to find/access and use the presented information. This outcome considers the three following domains: (1) how easy it was to find information, (2) how easy it was to understand the information (perception), and (3) whether the information was presented in a way that could be helpful for making an informed health decision. Participants will be asked to indicate the degree of agreement with six statements, measured using the 7-point Likert scale (1 being strongly disagree, 4 being neutral, and 7 being strongly agree). We will also include 1 open-ended question to get additional input on accessibility and usability. The outcome ‘overall accessibility/usability of information’ will be measured using the average of the 7-point Likert-type scale questions.

##### Satisfaction

We define satisfaction as participants’ impression of the recommendation’s presentation. We will ask participants about their level of satisfaction with different features of the format (e.g., order of information, length of document, etc.) [25]. Participants will be asked to indicate the degree of satisfaction with three questions measured using the original 7-point Likert-type scale (1 being very dissatisfied, 4 being neutral, and 7 being very satisfied). We will also include two open-ended questions to get their input on what they liked and disliked in the format.

##### Intended Behavior

We define intended behavior as participants’ intention in adopting and following the shared recommendation. We will ask participants if they have already followed the recommendation and have them respond with yes, no, unsure, prefer not to answer, or not applicable. Subsequently, we will ask how likely it is that they will follow the recommendation (if they haven’t already) and share the recommendation with others, through two questions measured using a 7-point Likert-type scale (1 being very unlikely, 4 being neutral, and 7 being very likely).

##### Preference

We define preference as a greater liking of one format over the other (PLR or SLV). After participants complete the tasks in the group they are randomized to, they will be asked to compare the PLR and the SLV formats. Participants will review the alternative format and indicate their preference for one of the two formats using a 7-point Likert-type scale (1 being strongly preferring the SLV, 4 being same preference for both, and 7 being strongly preferring PLR). Participants will also have the chance to explain their choice in a text box.

### Stratification

We will stratify our participants by region: Africa, Americas, Eastern Mediterranean, Europe, South-East Asia, and Western Pacific. We have not set a target sample per strata. The rationale behind stratification is to ensure an equal chance for participants from the same region to be randomized to either the intervention or control (as opposed to having all participants from Europe, for example, being randomized to the PLR format).

### Randomization

After stratification, participants will be randomly assigned in a 1:1 ratio to recommendation topic 1 or 2, then will be randomized again in a 1:1 ratio to the PLR or the SLV arms (see Figure 1). Randomization will ensure that recommendation formats will be equally distributed across participants to get balanced judgments on outcomes.

### Allocation Concealment

The allocation sequence is concealed using SurveyMonkey® software based on a commercial, but unknown algorithm without a pre-identified sequence.

### Blinding

Participants will know that the trials are testing different formats to present health recommendations but will be blinded with respect to what formats are being compared and to the group to which they are allocated. Participants will not be aware of their random allocation to the PLR or SLV format until disclosure. Thus, participants will be blinded for all outcomes except the secondary outcome of preference. We will blind data analysts involved in data interpretation and manuscript writing to reduce bias in data analysis and interpretation [26, 27]. To reduce bias, we will draft each population’s manuscript with general group labels (i.e., group A vs B) and agree on the final interpretation before the group allocation (optimized PLR or SLV) is revealed.

### Sample Size Calculation

The sample size was calculated using the primary outcome of understanding. For this two-sided (*α* = 0.05) superiority analysis, these computations were made based on a t-test with the null hypothesis that there is no difference between the PLR version and the SLV in understanding of information.

*H*_*0*_: PLR version = SLV

*H*_*1*_: PLR version ≠ SLV

We will determine whether we can reject the null hypothesis of no difference in understanding between the formats of presenting recommendations. We assume a correct response rate (regarding understanding of information) of 75% in the PLR arm and 60% in the control arm [relative risk =1.25] based on a recent RCT and prior trials on Plain Language Summaries (PLS) of summary of findings table [19, 24]. With typical alpha (0.05) and beta (0.8) parameters and an allocation ratio of 1:1, 240 participants (120 in each arm) would be required (Stata/SE 16.0) [28]. Assuming 15% non-completion, we will recruit 282 participants from each population and adjust this number if non-completion is higher.

Since we are using two different recommendation topics with each population, we might observe an interaction between the recommendation topic and our outcome of understanding. Thus, when we reach half of our intended sample size, we will conduct an interim analysis for a possible interaction effect. If the data suggests that there might be an interaction, we will consider modifying our initial sample size. This new sample size cannot be determined in advance since we cannot anticipate the magnitude of the potential interaction effect, but it will be determined based on published guidance on power calculations for credible subgroup effects [29]. If needed, we will submit an amendment that explains the new sample size and need for additional recruitment based on the magnitude of the interaction effect identified. In summary, an enhanced sample size, if indicated and feasible, will allow us to conduct more meaningful analysis by exploring the results by topic of recommendation.

### Consultation and Pilot Testing

We will use the same survey template for all populations; however, the “understanding” questions will be appropriately different for each recommendation. We pilot tested the surveys during the pre-trial engagement work to gather feedback from Canadian youth advisors, Cochrane Consumer Network, adult stakeholders, and parent stakeholders. We had 10 youth, 29 adults, and 5 parents pilot test the survey and provide feedback on the time to complete the survey, length of the survey, clarity of questions, internet difficulties, and any other comments to enhance the survey experience. Revisions were made to the surveys using pilot test feedback until no errors or inconsistencies were detected and the surveys were easily understood. The survey language used for each population is tailored based on the feedback from stakeholders. Pilot test participant results will not be included in the final analysis.

### Statistical Analysis

Each population’s results will be analyzed separately, and we will explore subgroup effects within each group. These subgroups are the topic of recommendation, health literacy level, and English proficiency level of participants. We will also pool the results across populations and conduct a meta-analysis for our primary outcome of “understanding” and other secondary outcomes. In the final analysis, after pooling the results, we will explore for potential interaction related to the recommendation topic and participants’ health literacy level. We will conduct the analyses using IBM SPSS® (Statistical Package for Social Sciences) version 23.

#### Descriptive Analysis

We will summarize participant baseline characteristics and outcomes using means and standard deviations (SD) for continuous variables, and proportions for categorical variables.

#### Inferential Analysis

We will perform a primary analysis including all randomized participants. We will exclude participants who take less than 6 minutes to complete the survey. For the outcome of understanding, we will use *χ*2 tests and risk difference with 95% CIs to compare the proportion of correct responses between groups. For the outcomes of accessibility and satisfaction, we will use t-tests and mean differences 95% confidence intervals (95% CIs) to compare means and SDs between the intervention and control groups. Finally, we will present preference as mean (SD) overall and for both trial arms. Skewness, Shapiro-Wilk tests, and histograms will be used to evaluate whether the distribution was shifted toward the same preference in both groups. Levene’s test of equal variances will be used for all t-tests and degrees of freedom will be adjusted for p < 0.05. We will report all p-values to three decimal places, with values less than 0.001 reported as < 0.001. We will consider statistical significance at p < 0.05. Furthermore, we will conduct a meta-analysis across the three trials by combining results of the three trials. Our a priori hypothesis is that there is no difference across the three populations despite the differences in population characteristics and slight difference in the presentation formats for each population. However, we will explore differences between trials applying the GRADE approach to evaluating inconsistency which includes the chi square test, the I^2^ value, overlap in confidence intervals and the differences in the point estimates of effect for all outcomes [30].

### Qualitative Methods

We will interview a sample of survey participants to explore their user experience with two recommendation formats. These semi-structured interviews will help us contextualize the results of the survey and understand participants preference for receiving the information.

#### Selection of Participants

At the end of each survey, participants will be invited to contribute to a virtual, one-on-one, semi-structured interview, conducted by the research coordinator for each leading site. Interested participants will voluntarily provide their email address at the end of the survey, and the research coordinator will follow-up with them through email.

#### Interviews

We will invite individuals from the list of participants who have agreed to participate in the qualitative interviews. For the parents’ population, we will use purposive sampling (inviting participants based on region, ethnicity, gender and topic of recommendation). For the adults and youth populations, we will not use a purposive sample, however, we will collect demographic data from the participants at the time of the interview. All interview participants will be compensated for their time with a gift card valued at $25 CAD.

Prior to the interview, participants will be provided with the consent form for the semi-structured interviews. Following that, a Zoom invite will be shared with the participant. Prior to the start of the interview, the research coordinator conducting the interview will obtain verbal consent from the participant for participating in the interview and for the interview to be audio-recorded. If the participant chooses to leave their video camera on during the interview, consent will be acquired for video-recording the interview. Consent will be documented on a log by the coordinator. If participants decline recording of the interviews, we will not move forward with the interview. We will transcribe interview recordings verbatim and de-identify all interview transcripts.

Interviews will be conducted in English by trained interviewers and will take approximately 30-60 minutes. Interviews will be guided by open-ended questions that cover the seven facets of the Honeycomb model to verify user’s experiences with the different recommendation formats: usefulness, usability, findability, accessibility, desirability, credibility, and value [31]. We will pilot test the interview guide within the three populations to refine the questions and language based on the populations’ unique needs prior to the interviews.

### Data management

All recordings will be stored on a password protected device and secure network and accessed only by members of the research team. The recordings will be kept for five years, at which point they will be destroyed. All transcribed recordings will be reviewed and verified by the interviewer or a second research team member prior to being de-identified. NVivo software (QSR, 2018) will be used for data coding and management.

### Data analysis

Qualitative data analysis will follow a hybrid inductive/deductive method [32]. This method allows for flexibility to utilize an established framework during analysis (deductive) as well as, identify codes that emerge from the data (inductive). The 7 elements of the honeycomb model for user experience (figure 4) informed the creation of the interview guide and were adapted and used as the initial model for analysis (see Appendix 2 for interview guide) [31]. The seven elements of the model will guide coding with inductive coding of data that extends beyond the model.

**Figure 4.**
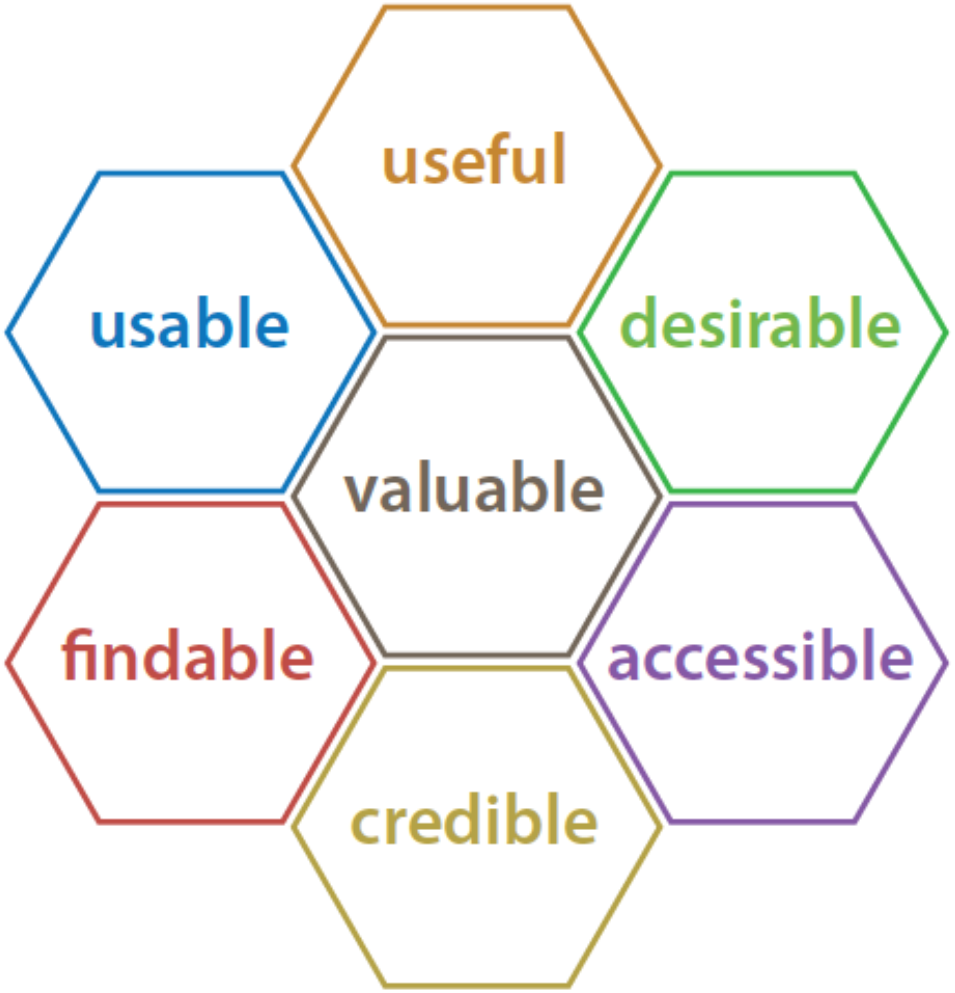
Honeycomb model of user experience

Data collection and analysis will occur iteratively [33]. The analysis will follow six steps: 1) Data cleaning-comparing the transcript with the recording and doing the cleaning to ensure alignment 2) generating initial codes using line by line coding 3) searching for themes guided by the honeycomb model. 4) Reviewing the themes and generating a “thematic map”, 5) defining and naming themes guided by the honeycomb model and 6) writing the report. Data collection will occur until saturation is reached [34].

We will develop joint displays, explicitly merging the results from the three populations’ data sets through a side-by-side comparison to assess how findings are aligned among the three data sets [35].

### Rigor

Strategies for maintaining rigor [36] will be used such as utilize a detailed study log and audit trail to ensure transparency. A coding system will be utilized and refined throughout the iterative data collection and analysis stages and each site will have a primary coder and a secondary coder to code approximately 10% of the transcripts and compare to maintain intrarater reliability [36]. We will use iterative member checking, reflecting back statements made during interviews to participants for clarity and understanding.

We will also use our trial survey data to triangulate our findings with survey responses. Our analysis team will reflect on and disclose our own digital experiences and consider this in our iterative analysis process.

## Reporting

We will report our quantitative findings according to the Consolidated Standards of Reporting Trials (CONSORT) reporting guidelines and qualitative findings following the Consolidated criteria for reporting qualitative research (COREQ) [37, 38]. For the quantitative results, we will pool the results across the three populations and conduct a meta-analysis for our primary outcome of “understanding” and other secondary outcomes. We will explore results for each population separately as well as compare results across populations.

Finally, we will crossmatch and compare the results of the qualitative interviews with those of the quantitative surveys and try to better understand and contextualize the RCT findings. Based on the results, we will discuss possible changes to the PLR format.

## Discussion

Guideline developers typically create recommendations for health professionals or health agencies [1]. However, particularly in the context of COVID-19 and any future pandemics or topics of high public interest, the general public must have access to health recommendations that they can understand and trust [39]. For example, public understanding and acceptance is required for implementation and uptake of many public health recommendations such as vaccination, using facial masks, and social distancing. Further, parents are making decisions for children who cannot make decisions for themselves, and they need trustworthy recommendations specific to this unique population (e.g., vaccine safety and efficacy for children). On the other hand, youth access health recommendations about COVID-19 and make independent decisions about implementing public health measures, like social distancing and face mask use [3].

It has become crucial to engage end-users in shaping behaviors and activities that will eventually affect them and their communities. Their involvement will improve the relevance, usefulness, and transparency of guidelines [40]. More importantly, it will ensure that these guideline products are understandable and acceptable. We believe that when the public is presented with plain language recommendations, they will have a better understanding of guideline recommendations. These trials are designed with the input of end-users from three populations: youth, adults, and parents. Results will inform the best ways to make recommendations understandable and accessible to the public, thus increasing the public’s confidence in science and evidence uptake.

## Supporting information

SPIRIT Checklist

Semi-structured interview guide

Survey Consent Form Sample

Interview Consent Form Sample

## Data Availability

Anonymous survey data will be publicly available from an open access online repository to be chosen within 2 years of data collection.

## Ethics and dissemination

After review, the Hamilton Integrated Research Ethics Board (HiREB) has approved this study. The University of Alberta Research Ethics Board has approved the parent portion of this study. The results of this study will be published in peer-reviewed journals: one for each population, and one presenting the pooled results. We also aim to present the results in national and international conferences and distribute them through other media. The authors of this protocol follow the ICJME authorship criteria.

## Availability of Data Materials

Anonymous survey data will be publicly available from an open access online repository to be chosen within two years of data collection.

## Funding and Sponsor

This work is supported by the “Canadian Institutes of Health Research” (CIHR), grant number: GA3-177732. McMaster University is the sponsor of the trials and can be contacted through the principal investigator, Dr. Holger Schünemann at.

CIHR had no role in study design; collection, management, analysis, and interpretation of data; writing of the report; and the decision to submit the report for publication.

Funder Contact Information:

*Canadian Institutes of Health Research*

*160 Elgin Street, Ottawa, ON, Canada*

*K1A 0W9*

*(613) 954-1968*

Sponsor Contact Information:

*Holger J. Schünemann*

*1280 Main St. W*., *Hamilton, ON, Canada*

*L8S 4K1*

*(905) 525-9140 × 24699*

## Consent to Participate

The study survey includes the consent form that describes the study purpose, confidentiality, potential risks, and voluntary participation (Appendix 3). Consent will be obtained from all participants in the study by overt action (i.e., by completing the survey). As for the interview, consent form will be shared via email (Appendix 4), and the research coordinator conducting the interview will obtain verbal consent from the participant for participating in the interview and for the interview to be audio-recorded.

## Data Protection and Confidentiality

Anonymized survey data will be stored in SurveyMonkey® password-protected software. Participants will be given the option to provide their names and email addresses in a separate survey link. Names and email addresses will be stored in a separate document on a secure password-protected computer and will not be linked to the survey responses. Only designated members of the research team will have access to this data.

## Authors’ Contributions

HJS, KP, LH, NJB, and MO are the principal investigators of the study and together with JM, EA, TK, DPR, SE, SDS and TL designed and established this research project. RC, LS, MM, SS, MP, and JR conducted pre-trial engagement work, piloted the survey and provided methodological input. RC, LS, MM and SS were responsible for drafting the ethics applications. RC and HJS were responsible for registering the protocol on the website, www.ClinicalTrials.gov. RC, LS, MM and SS are responsible for the coordination of the study. All authors participated in the writing and revision of the manuscript and approved its final version.

## Trial Registration

The trials were registered on Clinicaltrials.gov, ID: NCT05358990.

## Protocol Version

Version 1.4, August 08, 2022

## Additional Acknowledgements

Dr. Hartling is supported by a Canada Research Chair in Knowledge Synthesis and Translation. Dr. Scott is supported by a Canada Research Chair in Knowledge Translation in Child Health. Dr. Hartling and Dr. Scott are Distinguished Researchers with the Stollery Science Lab supported by the Stollery Children’s Hospital Foundation.

Dr Pottie is supported by the Ian McWhinney Research Chair in Family Medicine, Western University.

We gratefully acknowledge and thank the youth advisors, parents’ advisors and the adult Cochrane Consumer group who participated in workshops and meetings, provided feedback on study materials, and pilot tested the survey.

We gratefully acknowledge and thank Dr. Jozef Suvada and St. Elizabeth University in Slovakia who are helping us with recruitment efforts.

## Supplemental Material

Appendix 1. SPIRIT Checklist

Appendix 2. Semi-structured interview guide

Appendix 3. Survey Consent Form Sample

Appendix 4. Interview Consent Form Sample

