## Supplementary material for "Knowledge mobilization activities to support decision-making by youth, parents, and adults using a systematic and living map of evidence and recommendations on COVID-19: protocol for three randomized controlled trials and qualitative user-experience studies": Semi-structured interview guide

**Appendix 2- Semi-structured interview guide, sample questions**

After our informed verbal consent process is complete, we will start the interview process by showing the participant two formats of the COVID-19 recommendation which they saw in the survey. We will allow time for the participant to review the two formats and also give the participant time to ask any questions they may have.

We will then ask the participant our questions, for example:

For descriptive reasons about our interview sample, we would like to collect five additional data points if you are okay with sharing your responses with me:

1. What country do you currently reside in?
2. How comfortable are you with reading health information (on a scale 1-7, 7 being very comfortable and 1 being very uncomfortable)?
3. How would you identify your racial and ethnic background?
4. How would you describe your gender?
5. What is the highest level of education you have completed?
6. Which of the two formats did you see in the survey that you completed online?
7. What is your overall impression of each format, and why?
8. Which of the two formats do you prefer, format 1 or format 2, and why?
   a. *Probe*: Which format would you choose to share with your friends and family?
   b. *Probe*: What do you like about it? For example, document length, colors, language used, text size, etc.
   c. *Probe*: What would you change about this format to make it better?
9. Which format is easier to understand? Why?
   a. *Probe*: What are the most important features to make these formats useful and easy to use?
10. Was there any information you were looking for but did not find? If so, what was it?
11. Think about your understanding of the topic before and after reading the recommendation. Did the recommendation help you understand the information any better?
    a. *Probe*: What were takeaway/home messages for you from each format?
12. Would you use this Plain Language Recommendation format in the future to make health decisions?
13. Is there anything else you would like to share with me?

Would you like to receive a gift card valued at $25 CAD?

a. Yes
b. No

(Please note that the above answer will be recorded on the participant ID and verbal consent log)
