## Supplementary material for "Knowledge mobilization activities to support decision-making by youth, parents, and adults using a systematic and living map of evidence and recommendations on COVID-19: protocol for three randomized controlled trials and qualitative user-experience studies": Survey Consent Form Sample

**Appendix 3- Survey Consent Form**

**Study Title**: Knowledge mobilization activities to support decision-making by youth, parents and adults using a systematic and living map of evidence and recommendations on COVID-19

**Sponsor’s Study ID**: GA3-177732

**Study Doctor**: *insert name, department and telephone or pager number*

INTRODUCTION

*Describe any conflict of interest that exists or may appear to exist as it relates to any of the investigators, study staff or member of their immediate family. A conflict of interest exists if there is a potential benefit to the investigator(s), study staff or member of their immediate family beyond the professional benefit from academic achievement or presentation of the results. Examples include, but are not limited to, speaker’s fees, travel assistance, consultant fees, honoraria, gifts, and intellectual property rights such as patents. A declaration of conflict of interest should include the identity of the person with the conflict of interest, the type of incentive or inducement, and its source. See examples below.*

You are being invited to participate in a trial (a type of research study). You are invited to participate in this trial because you are a [*parent, youth or adult*] and we want to hear your feedback on the presentation of health recommendations related to COVID-19. This consent form provides you with information to help you make an informed choice about participation in the research. Please read this document. Please take your time in making your decision. You may find it helpful to discuss it with your friends and family.

Taking part in this study is voluntary. You have the option to not participate at all or you may choose to leave the study at any time. You will have the option to leave your name and email to enter a draw at the end of the survey for a chance to win [1 of 10 gift cards valued at $25 CAD, or 1 of 5 gift cards valued at $50 CAD.] Whatever you choose, it will not affect your relationship with McMaster University or [*insert name of recruitment site*].

IS THERE A CONFLICT OF INTEREST?

The researchers have no conflicts of interest to declare related to this study.

WHY IS THIS STUDY BEING DONE?

COVID-19 recommendations are usually developed for healthcare professionals (for example doctors, health organizations, etc.). We want to make sure that these recommendations can be used and understood by everyone. The purpose of this study is to make COVID-19 recommendations more accessible and easier for parents and caregivers, adults, and youth to understand. We are evaluating recommendations presented in two different formats and asking questions about understanding, satisfaction, and preference for the recommendation format.

HOW MANY PEOPLE WILL TAKE PART IN THIS STUDY?

It is anticipated that a total of 720 people will take part in this study from all around the world, which includes 240 adults, 240 youth and 240 parents. Study recruitment will take roughly three months to complete, and the results should be known two months after recruitment is completed.

WHAT WILL HAPPEN DURING THIS STUDY?

ASSIGNMENT TO A GROUP

If you decide to participate, then you will be randomly assigned to one of two groups as described below. Randomization means that you are put into a group by chance (like flipping a coin). There is no way to predict which group you will be assigned to.

WHAT IS THE STUDY INTERVENTION?

We are testing two different formats of COVID-19 health recommendations. You will be randomized to either the intervention or control and will receive one of the two formats. Each format will contain the same information; however, the presentation, language, and aesthetics will appear slightly different. Both groups will have to read the recommendation they receive.

WHAT ARE THE STUDY PROCEDURES?

Questionnaires

You will be asked to read and judge the format you have been assigned to. To do this, you will answer an online questionnaire which takes about 15-25 minutes to complete. The information you provide is for research purposes only.

Optional Research

At the end of the questionnaire, there will be an option to indicate whether you are interested in participating in a one-on-one interview to share your thoughts on the recommendation format with the researcher.

HOW LONG WILL PARTICIPANTS BE IN THE STUDY?

Your participation will take about 15-25 minutes until you complete the questionnaire.

CAN PARTICIPANTS CHOOSE TO LEAVE THE STUDY?

You can choose to end your participation in this research (called withdrawal) at any time without having to provide a reason. You can withdraw before you finish the questionnaire by closing your browser or navigating away from the survey. All collected responses are anonymous, so we cannot identify which responses were yours. Once you submit your response, we cannot remove or change it.

WHAT ARE THE RISKS OR HARMS OF PARTICIPATING IN THIS STUDY?

There are no foreseeable risks for participating in this study.

WHAT ARE THE BENEFITS OF PARTICIPATING IN THIS STUDY?

You may not benefit directly from participating in this study; however, your participation will help inform the research team which recommendation format is easier to understand for [youth, adults or parents]. This means that your participation will help make COVID-19 health recommendations more accessible to [adults, parents, or youth] all around the world.

HOW WILL PARTICIPANT INFORMATION BE KEPT CONFIDENTIAL?

**Note:** If there will be disclosure of personal identifiers, i.e., disclosed on any research-related information/documents including samples or scans, or as part of the unique identifier, these disclosures must be justified in the REB application and approved. Please ensure that you are aware of institutional and REB policies with respect to the disclosure of personal identifiers.

If you decide to participate in this study, the research team will only collect information they need for this study. Records identifying you at this center (ex. name and email address) will be collected by McMaster University and will only be shared with [*name of recruitment site*] if you requested to receive the trial results or would like to participate in an interview. These records will be kept confidential and, to the extent permitted by applicable laws, will not be disclosed or made publicly available, except as described in this consent document.

Authorized representatives of the following organizations may look at your original (identifiable) records at the site where these records are held, to check that the information collected for the study is correct and follows proper laws and guidelines.

- CIHR, the Sponsor of this study
- The research ethics board who oversees the ethical conduct of this study in Ontario
- This institution and affiliated sites, to oversee the conduct of research at this location

Information that is collected about you for the study (called study data) may also be sent to the organizations listed above. Representatives of Clinical Trials Ontario, a not-for-profit organization, may see study data that is sent to the research ethics board for this study. Your name and email address (if you choose to leave them) will not be used. The records received by these organizations may contain your participation code.

Studies involving humans sometimes collect information on race and ethnicity as well as other characteristics of individuals because these characteristics may influence how people perceive and favor different interventions. Providing information on your race or ethnic origin is optional.

Communication via e-mail is not absolutely secure. We do not recommend that you communicate sensitive personal information via e-mail.

If the results of this study are published, your identity will remain confidential. It is expected that the information collected during this study will be used in analyses and will be published/ presented to the scientific community at meetings and in journals. Even though the likelihood that someone may identify you from the study data is very small, it can never be completely eliminated.

WHAT IS THE COST TO PARTICIPANTS?

Participation in this study will not involve any additional costs to you or your private health care insurance.

ARE STUDY PARTICIPANTS PAID TO BE IN THIS STUDY?

*Each participating site must ensure that the information below matches the compensation/reimbursement provided at that site. Site specific differences must be reflected in the Centre Initial Application and the site-specific consent form.*

If you decide to participate in this study, you will have the option of being entered into a draw for a chance to win [1 of 10 gift cards valued at $25 CAD, or 1 of 5 gift cards valued at $50 CAD.] .

WHAT ARE THE RIGHTS OF PARTICIPANTS IN A RESEARCH STUDY?

You have the right to be informed of the results of this study once the entire study is complete. If you would like to be informed of the results of this study, please leave your email address at the end of the survey.

Your rights to privacy are legally protected by federal and provincial laws that require safeguards to ensure that your privacy is respected.

By completing this survey, you do not give up any of your legal rights against the researcher, sponsor or involved institutions for compensation, nor does this form relieve the researcher, sponsor or their agents of their legal and professional responsibilities.

WHOM DO PARTICIPANTS CONTACT FOR QUESTIONS?

If you have questions about taking part in this study, you can talk to the research team, or the person who is in charge of the study at this institution. That person is:

___________________________	_________________________

Name [*Coordinator at recruitment site*]	Email

If you have questions about your rights as a participant or about ethical issues related to this study, you can talk to someone who is not involved in the study at all. That person is:

Office of the Chair of the Hamilton Integrated Research Ethics Board

905-521-2100	ext.42013

____________________________	________________________

Name	Phone
