## Supplementary material for "Knowledge mobilization activities to support decision-making by youth, parents, and adults using a systematic and living map of evidence and recommendations on COVID-19: protocol for three randomized controlled trials and qualitative user-experience studies": Interview Consent Form Sample

**Appendix 4- Interview Consent Form**

**Study Title**: Knowledge mobilization activities to support decision-making by youth, parents and adults using a systematic and living map of evidence and recommendations on COVID-19

**Sponsor’s Study ID:** GA3-177732

**Study Doctor:** *insert name, department and telephone or pager number*

**INTRODUCTION**

You have recently participated in an online survey about different COVID-19 recommendation formats. Thank you for completing the survey! You are now invited to participate in an interview to share your feedback on these formats and your preference for different presentations of COVID-19 recommendations. This will help us understand the results of the survey and improve the way COVID-19 recommendations are developed for and shared with the public.

This consent form provides you with information to help you make an informed choice about participating in this interview, before giving verbal consent. Please read this document and ask any questions you may have. All your questions should be answered to your satisfaction before you decide whether to participate in this interview.

After you read the consent form, the researcher will ask you a few questions to obtain your verbal consent if you chose to continue with the interview. Taking part in this interview is voluntary. You have the option to not participate, or you may choose to stop the interview at any time. Whatever you choose, it will not affect your relationship with McMaster University or [*insert name of recruitment site*].

**IS THERE A CONFLICT OF INTEREST?**

*Describe any conflict of interest that exists or may appear to exist as it relates to any of the investigators, study staff or member of their immediate family. A conflict of interest exists if there is a potential benefit to the investigator(s), study staff or member of their immediate family beyond the professional benefit from academic achievement or presentation of the results. Examples include, but are not limited to, speaker’s fees, travel assistance, consultant fees, honoraria, gifts, and intellectual property rights such as patents. A declaration of conflict of interest should include the identity of the person with the conflict of interest, the type of incentive or inducement, and its source. See examples below.*

The researchers have no conflicts of interest to declare related to this study.

**WHY IS THIS STUDY BEING DONE?**

COVID-19 recommendations are usually developed for healthcare professionals (for example doctors, health organizations, etc.). We want to make sure that these recommendations can be used and understood by everyone. The purpose of this study is to make COVID-19 recommendations more accessible and easier for adults, parents, and youth to understand. We are evaluating recommendations presented in two different formats and asking questions about understanding, satisfaction, and preference for the recommendation format.

**HOW MANY PEOPLE WILL TAKE PART IN THIS STUDY?**

It is anticipated that around 54 people will take part in these interviews from around the world, including 18 adults, 18 parents and 18 youth.

**WHAT WILL HAPPEN DURING THIS STUDY?**

You will be asked to participate in a one-on-one interview with a researcher over Zoom. You will be asked to explain what recommendation format you prefer out of the two formats presented and provide additional insight into your preference.

With your consent, this interview will be audio recorded with the researcher. By participating in the interview, you are giving consent and allowing the researcher to audio record the interview as part of this research.

You will have the option to leave your camera on or turn it off during the Zoom call. If you choose to leave your camera on, the recording will capture your video and audio with your consent.

**HOW LONG WILL PARTICIPANTS BE IN THE STUDY?**

The interview will last from about 30 minutes to 60 minutes.

**CAN PARTICIPANTS CHOOSE TO LEAVE THE STUDY?**

You can choose to stop and end your participation in this interview (called withdrawal) at any time without having to provide a reason.

Participants can withdraw their interview data up to 24 hours after participation in the interview, before the transcription process occurs by contacting the study coordinator. However, once the transcripts are transcribed and de-identified, it will no longer be possible to separate the data from the data set.

**WHAT ARE THE RISKS OR HARMS OF PARTICIPATING IN THIS STUDY?**

There are no foreseeable risks for participating in this interview.

**WHAT ARE THE BENEFITS OF PARTICIPATING IN THIS STUDY?**

You may not benefit directly from participating in this study; however, your participation will help inform the research team which recommendation format is easier to understand for [youth, adults or parents]. This means that your participation will help make COVID-19 health recommendations more accessible to [adults, parents, or youth] all around the world.

**HOW WILL PARTICIPANT INFORMATION BE KEPT CONFIDENTIAL?**

*Note: If there will be disclosure of personal identifiers, i.e., disclosed on any research-related information/documents including samples or scans, or as part of the unique identifier, these disclosures must be justified in the REB application and approved. Please ensure that you are aware of institutional and REB policies with respect to the disclosure of personal identifiers.*

If you decide to participate in the interview, the research team will only collect information they need for this study. The recordings will be transcribed (turned into written records) and de-identified 24 hours after the interview. The interview recordings will be stored locally at the institution conducting the interview in a secure location and only viewed by members of the research team. The recordings will be destroyed after 5 years.

Records identifying you at this center will be kept confidential and, to the extent permitted by applicable laws, will not be disclosed or made publicly available, except as described in this consent document.

The interviews will be conducted over Zoom video conferencing software. A link to their privacy policy is located here [https://explore.zoom.us/en/privacy](https://explore.zoom.us/en/privacy/). Please note that while this service is approved for collecting data in this study there is a small risk with any platform such as this of data that is collected on external servers falling outside the control of the research team. Please talk to the researcher if you have any concerns.

Authorized representatives of the following organizations may look at your original records at the site where these records are held, to check that the information collected for the study is correct and follows proper laws and guidelines:

- CIHR, the Sponsor of this study
- The research ethics board who oversees the ethical conduct of this study in Ontario
- This institution and affiliated sites, to oversee the conduct of research at this location

Information that is collected about you for the study (called study data) may also be sent to the organizations listed above. Representatives of Clinical Trials Ontario, a not-for-profit organization, may see study data that is sent to the research ethics board for this study.  Your name and email address (if you choose to leave them) will not be used. The records received by these organizations may contain your participation code.

Communication via e-mail is not absolutely secure. We do not recommend that you communicate sensitive personal information via e-mail.

If the results of this study are published, your identity will remain confidential. It is expected that the information collected during this study will be used in analyses and will be published/ presented to the scientific community at meetings and in journals. Even though the likelihood that someone may identify you from the study data is very small, it can never be completely eliminated.

**ARE STUDY PARTICIPANTS PAID TO BE IN THIS STUDY?**

*Each participating site must ensure that the information below matches the compensation/reimbursement provided at that site. Site specific differences must be reflected in the Centre Initial Application and the site-specific consent form****.***

To thank you for participating, we will be compensated for your time with an electronic gift card valued at $25 CAD (if you wish to receive it).

**WHAT ARE THE RIGHTS OF PARTICIPANTS IN A RESEARCH STUDY?**

You have the right to be informed of the results of this study once it is complete. If you would like to receive the study results, please share your email address with the researcher at the beginning of your interview.

Your rights to privacy are legally protected by Canadian federal and provincial laws that require safeguards to ensure that your privacy is respected.

**WHOM DO PARTICIPANTS CONTACT FOR QUESTIONS?**

If you have questions about taking part in this study, you can talk to the research team, or the person who is in charge of the study at this institution. That person is:

___________________________	_________________________

Name [*Research coordinator at recruitment site*]	**Email**

If you have questions about your rights as a participant or about ethical issues related to this study, you can talk to someone who is not involved in the study at all. That person is:

Office of the Chair of the Hamilton Integrated Research Ethics Board

905-521-2100 ext. 42013

__________________________	________________________

**Name**	**Phone**
